# A comparison of health care worker-collected foam and polyester nasal swabs in convalescent COVID-19 patients

**DOI:** 10.1101/2020.04.28.20083055

**Authors:** Brian Hart, Yuan-Po Tu, Rachel Jennings, Prateek Verma, Leah R Padgett, Douglas Rains, Deneen Vojta, Ethan M Berke

## Abstract

**Background:** The exponential growth of COVID-19 cases and testing has created supply shortages at various points in the testing workflow. As of April 15, 2020 FDA recommendations only allowed for the use of nasopharyngeal, flocked mid turbinate, or foam nasal swabs, all of which are in very low supply. Polyester swabs are more readily available and mass producible. We compare the performance of polyester and foam swabs stored in different transport media.

**Methods:** Both polyester and foam nasal swabs were collected from convalescent COVID-19 patients at a single visit. Using the foam nasal swabs as the comparator, sensitivity of the polyester swabs in each media were calculated, three by three tables were constructed to measure concordance, and cycle threshold (Ct) values were compared.

**Findings:** 126 visits had polyester and foam swabs stored in viral transport media (VTM), 51 had polyester and foam swabs stored in saline, and 63 had a foam swab in VTM and a polyester swab stored in a dry tube. Using nasal foam swabs as a comparator, polyester nasal swabs had a sensitivity of 86·5% when both samples were stored in VTM, 86·7% when both samples were stored in saline, and 72·4% when the polyester swab was stored dry and the foam swab was stored in VTM. Polyester and foam Ct values from the same visit were correlated, but polyester swabs showed decreased performance for cases with a viral load near the detection threshold and higher Ct values on average.

**Interpretation:** Polyester nasal swabs showed a reduction in performance from foam nasal swabs, but may still provide a viable sample collection method given the current supply shortages and public health emergency.

**Funding:** Laboratory testing was conducted with financial support from Thermo Fisher Scientific.

## Introduction

In the few months since the Centers for Disease Control and Prevention began tracking cases of coronavirus disease 2019 (COVID-19), more than 800,000 people have officially tested positive in the United States alone, resulting in more than 44,000 deaths^1^. The exponential spread of SARS-CoV-2 has resulted in an enormous demand for testing for the SARS-CoV-2 virus that causes the disease. This increased demand has put a strain on every level of the system from personal protective equipment (PPE) worn by health care workers while administering the test, to swabs for collecting the samples, to viral transport media (VTM) to store the sample until tested, to laboratory capacity^2,3,4^. Efforts to provide viable alternatives for each of these supply shortages are crucial to keeping the healthcare workforce safe and the testing throughput high.

The shortages of PPE and swabs are driven, in part, because the originally recommended nasopharyngeal (NP) test cannot be self-administered, requiring a change in PPE for each test. A recent study showed that patient-collected foam nasal swabs were comparable to health care worker-collected NP swabs for detecting SARS-CoV-2 virus, providing a safer and less invasive sampling method^5^. These results validate findings for influenza testing^6,7^. While these results open the possibility of using patient-collected foam nasal swabs for sample collection, reducing the risk of viral exposure for health care workers, several issues remain. In the United States, foam nasal and nylon flocked swabs are not as readily available or mass produced as polyester swabs^8^. Additionally, swabs are typically stored and transported in VTM under refrigeration at or below 4°C. With VTM and swab supplies running low, and difficulties in obtaining sufficient refrigeration space for the massive number of samples arriving at the labs to be tested, testing requirements must be reevaluated to see if they can be safely altered^9,10^. Recent work has shown that saline may be a suitable replacement for VTM^11^. Since these findings were released, the Food and Drug Administration (FDA) has updated their testing recommendations to allow for a wider variety of substances to be used for viral transport and stabilization and allow for self-collected foam nasal swabs when VTM and NP swabs are not available^12^.

To address the dwindling supply of recommended swabs, we compared the relative performance of polyester and foam nasal swabs for detecting SARS-CoV-2, stored and transported either in VTM, saline, or in a dry tube.

## Methods

### Population and Sample Collection

Patients who have previously tested positive for SARS-CoV-2 at any site from the original study in Washington state were approached to return for additional testing^5^. A cohort of 63 positive patients returned 7–9, 14–18, and 28–31 days after their initial positive diagnosis for longitudinal sample collection, with swabbing continuing until all results for samples from a single visit were negative. A second cohort consisted of participants who also previously tested positive. They were contacted by ambulatory clinic staff and asked to return for a single visit as soon as possible.

People who agreed to participate in either study were consented by medical staff. Participants were evaluated at a designated ambulatory clinic site that only saw patients that were confirmed with a prior positive SARS-CoV-2 reverse transcription polymerase chain reaction (RT-PCR) test. Inclusion criteria included a previous positive SARS-CoV-2 test and the ability to consent and agree to participate in the study. People who were not able to demonstrate understanding of the study, not willing to commit to having all samples collected, had a history of nosebleed in the past 24 hours, nasal surgery in the past two weeks, chemotherapy treatment with documented low platelet and low white blood cell counts, or acute facial trauma were excluded from the study. Health care workers used a written consent form to explain the study and give eligible patients the opportunity to decline. This study protocol was deemed to be part of a minimal risk research protocol by the Office of Human Research Affairs at UnitedHealth Group.

In the first cohort of 63 SARS-CoV-2 positive individuals, three nasal swabs were collected by a health care worker from each patient: a foam swab and two polyester swabs. Patients born in an odd numbered year first had their left nostril swabbed by the foam swab followed by their right nostril being swabbed by the two polyester swabs. Patients born in an even numbered year had their right nostril swabbed by the foam swab followed by their left nostril being swabbed by the two polyester swabs. The first of the two polyester swabs was stored in VTM and the second stored in a dry plastic tube without VTM. The foam swab was always stored in VTM. The two swabs stored in VTM were refrigerated and sent to a reference laboratory for immediate testing, while the dry polyester swab was stored without refrigeration for four days prior to testing. Due to participant testing constraints, the polyester swab to be stored in a dry tube was not collected from all patients. These swabs will be referred to as the foam, VTM polyester, and dry polyester swabs.

Patients in the second cohort were also convalescing from COVID-19. They were recruited immediately after their initial positive test result for SARS-CoV-2. During a single visit (between two and seven days after initial diagnosis), these patients were swabbed twice: once with a polyester swab and once with a foam swab. The order of sample collection was randomized in the same manner as in the first cohort. Both samples were refrigerated at 4°C after storage and tested in a reference laboratory as soon as possible.

All nasal swab samples were collected by health care workers in the following manner: 1) gently insert the swab in the vertical position into one nasal passage until there is gentle resistance 2) rotate the swab around the nasal vestibule for 10-15 seconds, 3) repeat in the other nostril, and 3) break off the head of the swab in the storage tube. All samples were marked with a patient study number, the swab type, and the media type if any.

RT-PCR was performed by a reference laboratory using the TaqPath™ COVID-19 Combo Kit (ThermoFisher; A47814). Cycle threshold (Ct) values for all samples were reported back to the clinical sites based on three different RNA targets: S Gene, ORF1ab, and N Gene. A higher Ct value corresponds to a lower viral load. Per the reference laboratory standard protocols, a sample was considered positive if two or three Ct values were less than or equal to 37, negative if all three Ct values were greater than 37, and inconclusive otherwise. All inconclusive samples were retested and the results of the retest were used, per manufacturer protocol. While a Ct value of 37 was used to determine the clinical results of the test, RT-PCR continued until 40 cycles were completed. Therefore, the maximum Ct value in the data is 40. Foam swabs were stored in 1 mL of VTM or 2 mL of saline and vortexed for three to five seconds. Polyester swabs were stored in 3 mL of VTM or 2 mL of saline and vortexed for three to five seconds. Dry polyester swabs were eluted in 1 mL of phosphate-buffered saline and vortexed for 30 seconds followed by a ten minute incubation. Ct values were normalized to estimated 3 mL Ct values to account for the difference in volumes. Specifically, 1Ct was subtracted from all results stored in 2 mL of saline or VTM, and 1·585 Ct was subtracted from polyester results stored in 1 mL. The Ct adjustments assume 90–100% PCR efficiency and follow the formula 2^−^*^dCt^ = F*, where dCt is the necessary Ct adjustment and F is the fold increase in volume.

### Statistical Analysis

We report the 3×3 confusion matrix showing the results of the polyester swabs against the results of the foam swabs. We estimate the sensitivity of the polyester swabs using the foam nasal swab as the comparator and report the 95% confidence intervals for the sensitivity estimates. For the purposes of sensitivity calculations, inconclusive results were counted as negatives and foam swabs were used as the comparator group. For each RNA target, we calculated the correlation of the normalized Ct values between the foam and polyester swabs for all pairs where at least one of the Ct values was < 40, indicating enough of the RNA target was detected to stop the RT-PCR machine short of the maximum Ct value. For the same subset of polyester/foam swab pairs with at least one Ct < 40 we also took the difference between the two Ct values and plotted the results in a box plot. Positive difference values indicate a higher Ct (and thus lower viral load) in the polyester samples and negative values indicate a higher Ct in the foam samples. All statistical analysis was performed using R version 3.6.1^13^.

## Results

Of the 63 patients in the first cohort, 29 were positive at the 7-9 day visit, five were positive at the 14–18 day visit, and zero were positive at the 28-31 day visit. One patient’s swabs from the 28-31 day visit leaked in transport and were not tested. The second cohort consisted of 76 participants, of which 51 had their swab samples stored in saline and 26 had their swab samples stored in VTM.

Polyester nasal swabs detected four fewer positive cases than foam swabs and had an estimated sensitivity (95% confidence interval) of 86·5% (77·3% - 95·8%) when stored in VTM (Table 1). Additionally, there were eight visits with an inconclusive VTM foam swab and a negative VTM polyester swab, but no visits with an inconclusive VTM polyester swab and a negative VTM foam swab. In saline, the polyester swabs detected two fewer positive cases and had an estimated sensitivity of 86·7% (74·5% - 98·8%) (Table 2). The dry polyester swabs detected five fewer cases than foam swabs in VTM and had an estimated sensitivity of 72·4% (56·1% - 88·7%) (Table 3).

**Table 1:** A 3×3 table of the test results for the foam and polyester nasal samples, both stored in VTM and tested immediately.

| Sensitivity (95% CI):<br>86·5% (77·3%, 95·8%) |  | VTM Polyester Nasal |  |  | Total |
| --- | --- | --- | --- | --- | --- |
|  |  | Positive | Negative | Inconclusive |  |
| Foam Nasal<br>in VTM | Positive | 45 | 6 | 1 | 52 |
|  | Negative | 3 | 63 | 0 | 66 |
|  | Inconclusive | 0 | 8 | 0 | 8 |
|  | Total | 48 | 77 | 1 | 126 |

**Table 2:** A 3×3 table of the test results for the foam and polyester nasal samples, both stored in saline and tested immediately.

| Sensitivity (95% CI):<br>86.7% (74.5%, 98.8%) |  | Saline Polyester Nasal |  |  | Total |
| --- | --- | --- | --- | --- | --- |
|  |  | Positive | Negative | Inconclusive |  |
| Foam Nasal<br>in Saline | Positive | 26 | 4 | 0 | 30 |
|  | Negative | 2 | 17 | 1 | 20 |
|  | Inconclusive | 0 | 1 | 0 | 1 |
|  | Total | 28 | 22 | 1 | 51 |

**Table 3:** A 3×3 table of the test results for the foam nasal swabs stored in VTM and tested immediately compared to polyester nasal samples stored in dry tubes without refrigeration for four days before being tested.

| Sensitivity (95% CI):<br>72.4% (56.1%, 88.7%) |  | Dry Polyester Nasal |  |  | Total |
| --- | --- | --- | --- | --- | --- |
|  |  | Positive | Negative | Inconclusive |  |
| Foam Nasal<br>in VTM | Positive | 21 | 5 | 3 | 29 |
|  | Negative | 2 | 26 | 2 | 30 |
|  | Inconclusive | 1 | 3 | 0 | 4 |
|  | Total | 24 | 34 | 5 | 63 |

Correlations between foam and polyester Ct values were highest in the saline samples, followed by the VTM samples and then the dry polyester versus VTM foam samples (Figure 1). While Ct correlations were highest in saline, the foam Ct value was lower than the polyester Ct value more than 63% of the time for the saline swabs, indicating the increased viral loads estimated using foam swabs (Figure 2). Similar biases toward lower Ct values appeared in the VTM polyester versus VTM foam and dry polyester versus VTM foam comparisons. The tendency toward lower Ct values for foam swabs is also apparent in the paired Ct plots (Figure 3). Figure 3 is further broken out by samples taken less than ten days from symptom onset and samples taken at least 10 days from symptom onset in Supplemental Figures 1 and 2.

**Figure 1:**
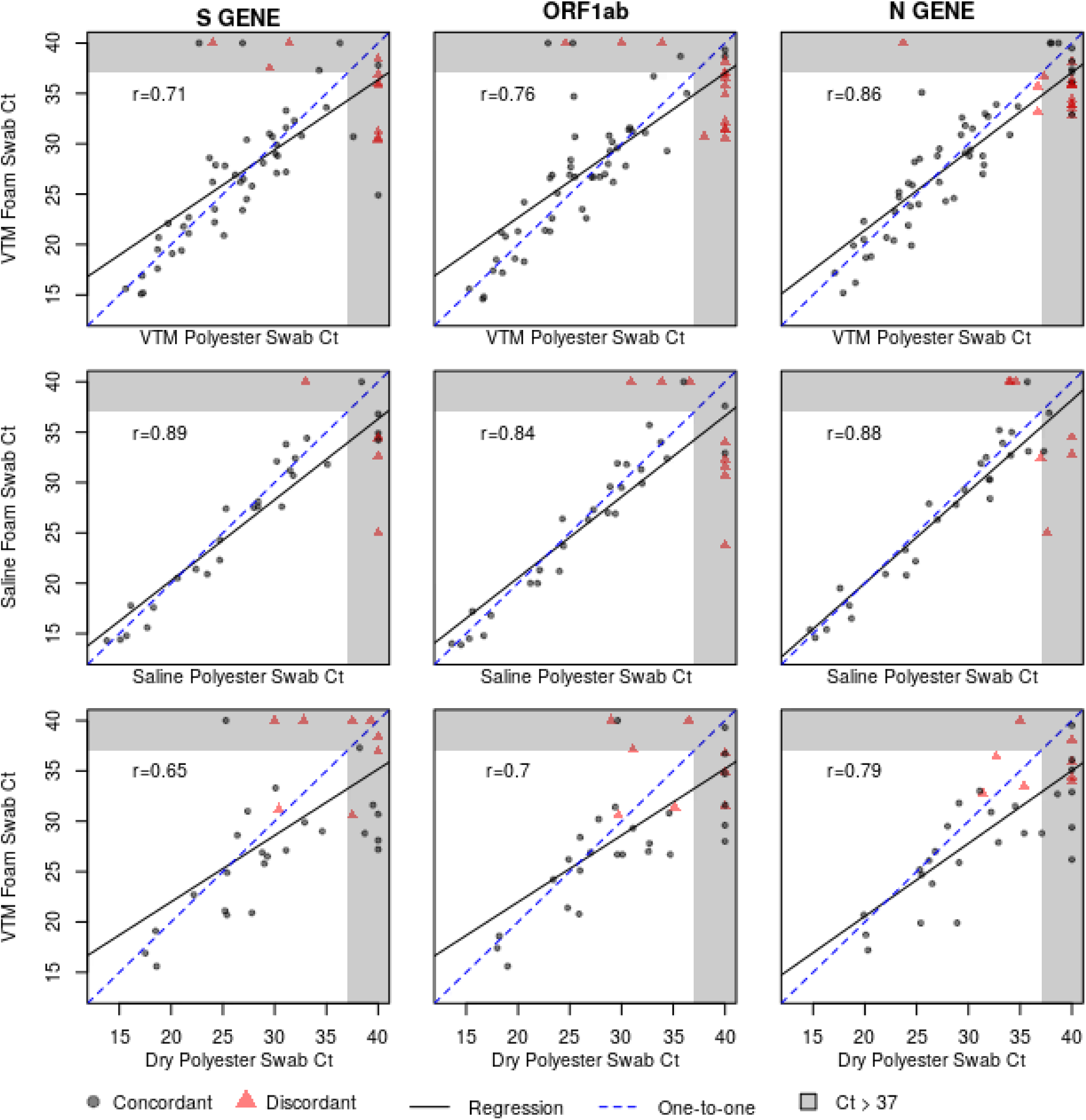
Plots showing the cycle threshold (Ct) values for each of the three RNA targets and three transport media. The black line represents the best fitting linear regression, the dashed blue line represents a perfect one-to-one relationship.

**Figure 2:**
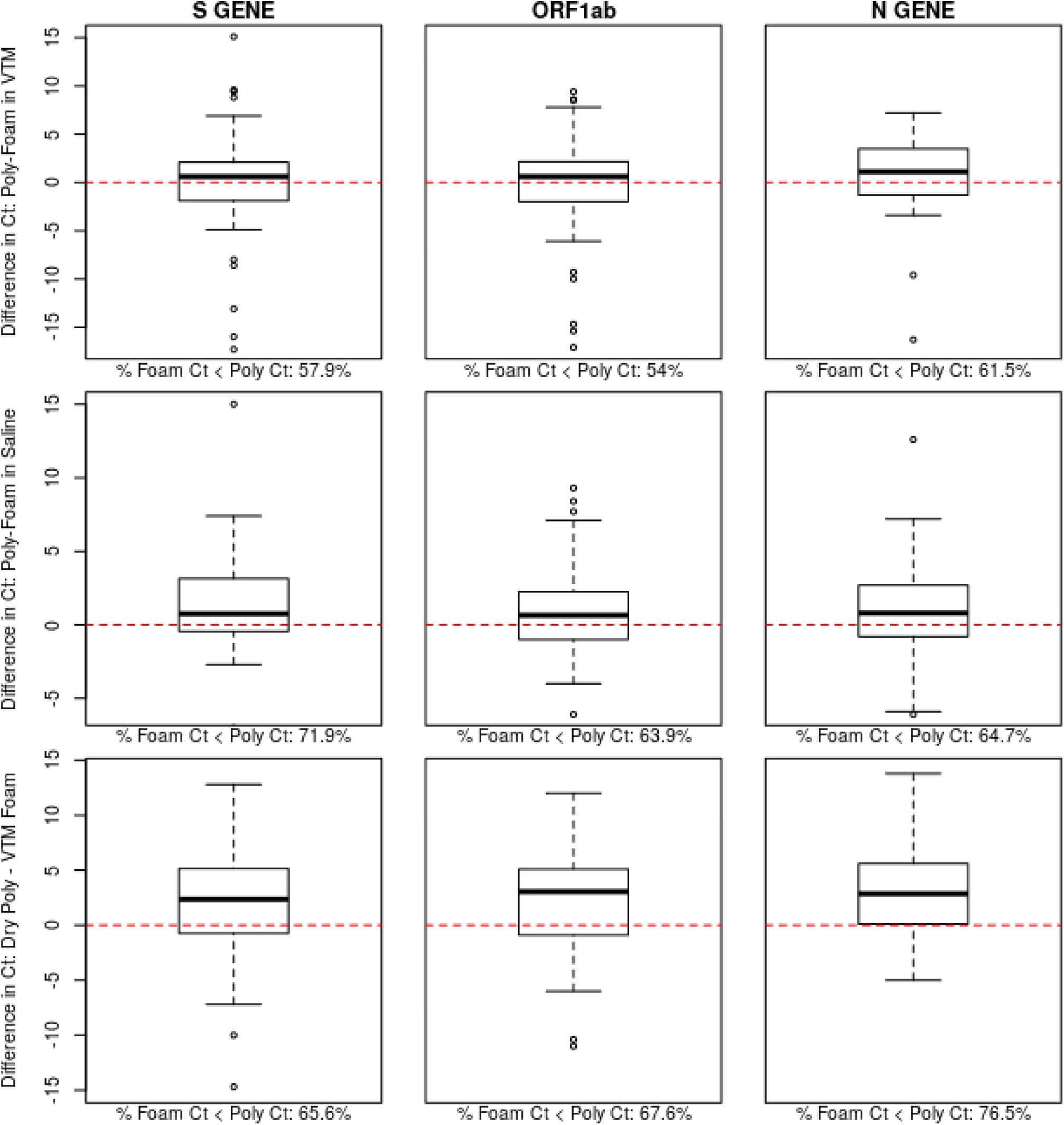
Plots showing the difference in cycle threshold (Ct) of the polyester and foam swabs collected at the same visits. Positive values represent higher Ct values in the polyester swab. The dashed red line represents equivalent Ct values. The percentage of samples for which the foam swab has a lower Ct value is shown below each sub-plot.

**Figure 3:**
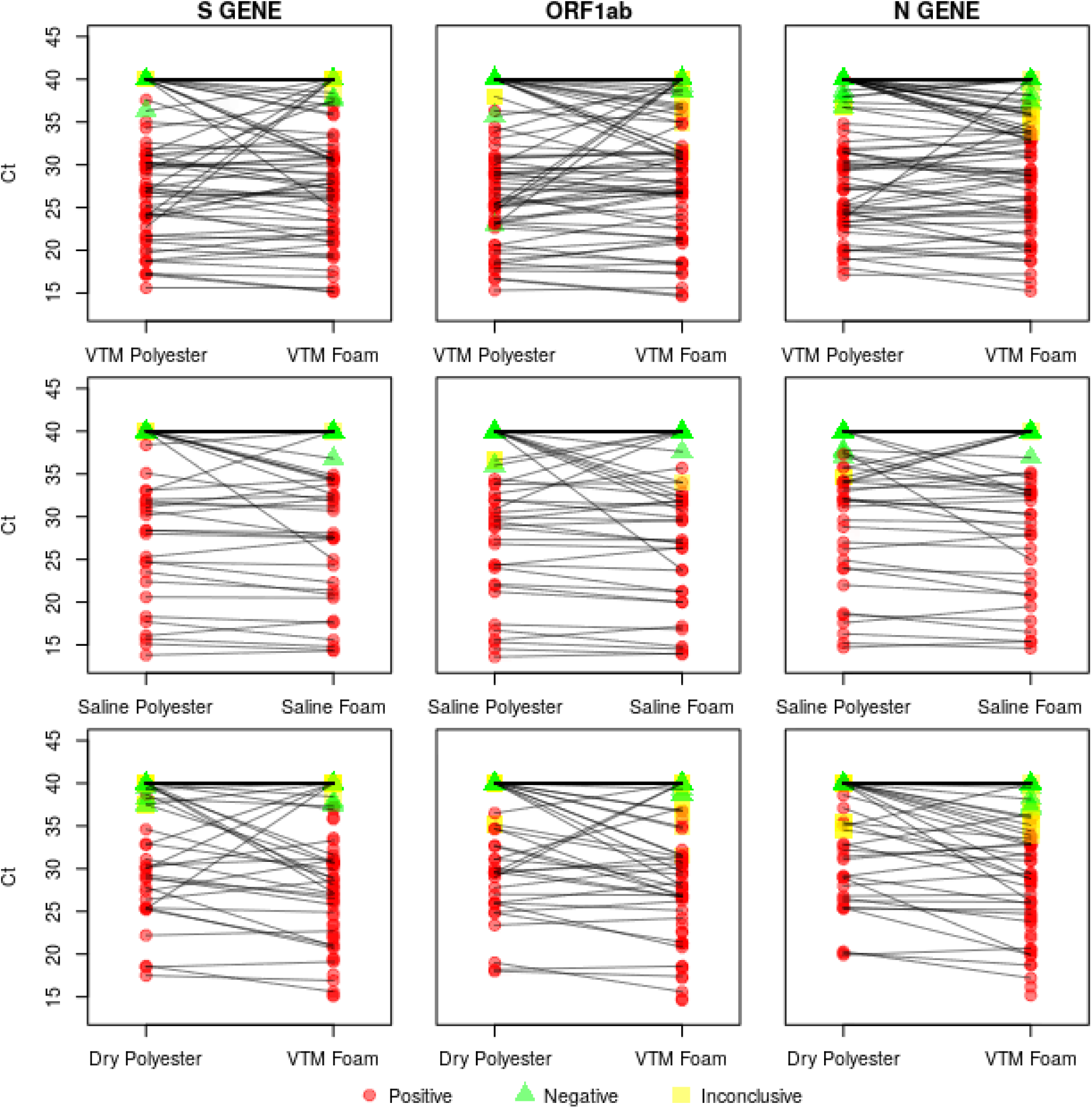
Paired Ct plots showing the polyester and foam Ct values for each transport media and target gene combination considered. Swabs collected at the same visit are connected by a black line.

## Discussion

Direct comparison of foam and polyester nasal swabs stored in VTM, in saline, or dry demonstrated decreased, but potentially adequate performance of polyester swabs. The estimated sensitivity of the polyester swabs was 86·5% and 86·7% in VTM and saline respectively. These calculations were conservative because they counted inconclusive polyester results as negatives for the purposes of sensitivity estimation, although no positive VTM foam swabs had a corresponding inconclusive VTM polyester swab. Additionally, foam nasal swabs do not represent the gold standard testing method, and the sensitivity calculations do not account for cases where the foam swab returned a positive result but the polyester swab did not. It is reasonable to believe these are indeed true positives when viewed in the light that all patients had previously tested positive for SARS-CoV-2.

The comparison of polyester and foam swabs did not differ between saline and VTM storage. While the estimated sensitivity above 86% may be deemed sufficient in times of a public health emergency, the strength of the findings is diminished for cases with a viral load near the positive/negative threshold of 37 cycles. This issue can be most clearly seen via the cluster of points along the upper edge of the right hand side of the S Gene and N Gene correlation plots. These points represent cases where foam swabs detected virus and the polyester did not. This finding is also reflected in the eight previously mentioned inconclusive VTM foam results that were negative for the VTM polyester swab. All three polyester versus foam swab comparisons exhibited a tendency for the foam swab to have lower Ct values than the polyester swab, indicating the foam swab’s superior ability to detect virus. Despite these reductions in performance, polyester swabs in VTM or saline may be an adequate sample collection method in cases where foam nasal and NP swabs have been entirely exhausted, a situation which exists in many locations.

The dry polyester swabs showed poor performance, but were put at a disadvantage by the study design. The degradation in performance of the dry swabs could arise from the lack of transport media, lack of refrigearation, extended time before testing, decreased sample collection due to it being the third swab in a nostril, or a combination of these factors. Dry swabs were only collected for the first follow-up visit in the cohort of 63 patients that also had swabs stored in VTM, resulting in a smaller sample size and different time since symptom onset than the other comparisons. Dry swabs have been found to have high sensitivity in other diagnostic settings and should not be ruled out entirely based on this study^14^.

The current study has several limitations. All participants were convalescing COVID-19 patients, and the time from first symptoms and first diagnosis to test date varied from test to test. As patients progressed further from their diagnosis date their viral loads dropped, creating more cases where the Ct values were near the border of detection. Other research has suggested viral load may have already peaked at the time of diagnosis followed by a slow decline over time^15^. The samples from this study may not be representative of testing in newly infected patients who are seeking their first SARS-CoV-2 test. Additionally, because no NP swabs were obtained for this study, performance of the polyester swab cannot be directly compared to the FDA’s preferred swabbing method^12^. Although final Ct values were adjusted for varying amounts of transport media, imprecision in these adjustments could alter the VTM polyester versus VTM foam comparison. Finally, the study was not designed to compare the performance of VTM and saline or to estimate an interaction of swab type and transport media type.

Despite these limitations, polyester swabs stored in VTM or saline may be a viable sample collection method for COVID-19 testing, especially in light of the shortages of other swab types. The viability of polyester swabs is most clearly demonstrated via the high correlation between polyester and foam Ct values from the same visit. Any recommendation for polyester swab usage should bear in mind that the decrease in performance near the border of detection may lead to false negatives in patients with low viral loads.

### Data Sharing Statement

A deidentified dataset will be provided upon reasonable request to the corresponding author.

## Data Availability

A deidentified dataset will be provided upon reasonable request to the corresponding author.

## ACKNOWLEDGMENTS

The authors acknowledge the contributions of James S Elliott and Lauren Kennington from Quantigen, Lindsay Nelson from UnitedHealth Group, and Garrett Galbreath, the health care workers, and staff from The Everett Clinic.

## Notes

### Competing Interest Statement

Dr. Leah Padgett and Douglas Rains perform contract work for Thermo Fisher Scientific. Dr. Tu is the principal investigator of a validation study for an Abbott point-of-care testing method.

### Funding Statement

Lab testing services for this study were funded by Thermo Fisher Scientific.

